# Building Resilience and Compassion in Conflict-Affected Communities: A School-Based Phenomenological Study in Colombia

**DOI:** 10.1101/2025.08.19.25333699

**Authors:** Lina Maria González-Ballesteros, Camila Andrea Castellanos-Roncancio, Oscar Eduardo Gómez Cárdenas, Isabela Osorio Jaramillo, Karen Ariza-Salazar, Luis Eduardo Mojica Ospina, Jennifer Clavijo Marin

## Abstract

In low- and middle-income countries such as Colombia, the increasing prevalence of mental disorders in youth has driven interest in school-based community-based psychosocial interventions aimed at prevention and wellness promotion. However, little is yet known about how key capacities such as resilience and compassion are perceived in these contexts, where patterns of access to resources are limited and cultural dynamics may differ significantly from those documented in Western, educated, industrialized, wealthy, and democratic countries from which most of the existing research originates. This study explored perceptions of resilience and compassion among children, adolescents and teachers in educational institutions in three departments of Colombia.

A qualitative observational study with a phenomenological approach was conducted within the framework of the intervention “Conmigo, Contigo y Con Todos – 3Cs” (with me, with you, and with everyone) which analyzed 833 focus groups: 166 with teachers and 667 with students in three measurement moments. The findings reveal that resilience and compassion are dynamic capacities shaped by emotional experiences and sociocultural context. The 3Cs Model facilitated processes of self-reflection, emotional regulation, reinforcement of mutual care, a deeper sense of community, and a reconfiguration of the school environment as a space for collective care and emotional safety. In addition, there was evidence of a re-signification of suffering among both teachers and students. These results underscore the value of contextually relevant, community-based, school-centered interventions for promoting mental health awareness and enhancing socio-emotional competencies in educational settings affected by adversity.

## Introduction

In Colombia, as in many other low- and middle-income countries, the growing prevalence of mental health disorders such as anxiety and depression among children and adolescents is increasingly concerning (Kohn et al., 2018; Ministry of Health and Social Protection & Colciencias, 2015; World Health Organization, 2021). This situation is exacerbated by structural social and cultural conditions that sustain the burden of mental illness and deepen existing treatment gaps, including exposure to armed conflict, substance use and abuse, and structural barriers to accessing health and education services. These same conditions often expose individuals to poverty, violence, and social exclusion, further aggravating mental health outcomes (Lund et al., 2010; Marroquín Rivera et al., 2020; Tol et al., 2013a; Yatham et al., 2018).

School-based, community-driven mental health interventions have emerged as essential strategies not only for prevention and the promotion of well-being, but also for fostering community transformation in settings affected by social exclusion and conflict (Bertsia & Poulou, 2023; Larson et al., 2020; Matos et al., 2022). These initiatives aim to enhance mental health literacy, encourage help-seeking behavior, reduce stigma, and strengthen local capacities for collective care and psychosocial support (Campos et al., 2018; Ma, Burn et al., 2023; Nobre et al., 2021). For example, school-based psychosocial programs that focused on improving students’ ability to recognize signs and symptoms of mental disorders were more likely to yield statistically significant gains, suggesting that targeting symptom recognition may be a particularly effective strategy for enhancing mental health knowledge among adolescents (Ma, Burn et al., 2023). More recently, such programs have also incorporated the development of socio-emotional skills such as resilience and compassion, recognized as key protective factors that support both individual well-being and the capacity for collective resilience in communities affected by adversity (Nobre et al., 2022).

Resilience has been conceptualized as the ability to overcome adversity (Bertsia & Poulou, 2023; Bonanno, 2004), maintaining psychological well-being in the face of adversity (Anderson & Priebe, 2021). It entails the activation of personal and social resources that enable individuals and communities to withstand, recover from, or transform adverse experiences. Similarly, compassion described as the act of being sensitive to the suffering of others (Peters & Calvo, 2014); has been associated with reduced levels of anxiety and depression, and with improved overall mental health and well-being (Avendaño-Vásquez et al., 2021; Dray et al., 2017; Gilbert, 2019, 2020; Lara-Cabrera et al., 2021; Windle, 2011). Beyond its clinical relevance, however, compassion must also be understood as a deeply cultural and communal phenomenon. (Goetz, Keltner, & Simon-Thomas, 2010) argue that compassion is a complex emotion with evolutionary roots that support cooperation, caregiving, and social cohesion. Although the capacity for compassion appears to be a universal human trait, its expression, valuation, and cultivation are significantly shaped by cultural norms, linguistic distinctions, emotional display rules, and contextual expectations. These cultural dynamics frame compassion not only as an individual disposition but as a socially embedded resource that sustains connectedness, solidarity, and communal resilience.

Together, these skills foster independence, responsibility, optimism, and social competence in students (Matos et al., 2022; Mullen et al., 2021; Srikala & Kishore Kumar, 2010); as well as contributing to more positive school environments, enhanced academic performance, and stronger educator engagement (Bertsia & Poulou, 2023; Bluth et al., 2015; Bluth & Blanton, 2014; Felver et al., 2016; Pepping et al., 2016).

Although the relationship between resilience and mental health has been widely studied, research exploring perceptions of resilience and compassion in Latin American populations remains limited (World Health Organization, 2021). This gap is particularly evident given the unique cultural dynamics and restricted access to mental health resources in the region, which differ markedly from those in Western, Educated, Industrialized, Rich, and Democratic (WEIRD) countries—contexts from which much of the existing evidence is drawn (Beyebach et al., 2021; Henrich et al., 2010). Understanding local perceptions is critical for guiding mental health policy and designing culturally responsive literacy programs that align with community needs and values (Roundtable on Health Literacy. Board on Population Health and Public Health Practice & Institute of Medicine, 2013).

Community psychology operates through a value-based praxis that connects reflection, research, and social action, fostering personal, collective, and relational well-being by emphasizing relational justice, collective agency, and meaningful community participation grounded in locally expressed needs and ideals (Prilleltensky, 2001). While the field articulates a clear commitment to social justice, it continues to grapple with aligning its stated values and actual practices. Advancing a socially just community psychology requires ongoing critical self-reflection, collective action, and the strategic engagement of professionals committed to transforming dominant institutional cultures through advocacy and structural change (Evans, Rosen, & Nelson, 2014). This alignment demonstrates how community-based action can foster relational justice and collective agency to support socio-emotional development in contexts of adversity.

In this context, the present study aimed to examine changes in perceptions of resilience and compassion among children, adolescents, and their teachers in schools located in three socioeconomically vulnerable Colombian departments affected by the armed conflict.

## Methods

### Study Design

A phenomenological observational qualitative qualitative study was conducted between June 2023 and December 2024 to explore changes in perceptions of resilience and compassion among children, adolescents, and teachers. This design provided an in-depth understanding of the subjective experiences, behaviors, and beliefs of individuals and communities within their natural educational contexts (Kumar Das, 2022).

### Participants

Forty-six of the 56 public educational institutions in the municipalities of Vaupés, Amazonas and Boyacá were selected based on criteria defined by the Secretaries of Education and considering the public order and political situation. These were randomly assigned in equal parts to the intervention (n=23) and control (n=23) groups. Theoretical sampling by maximum variation was used, organizing three focus groups of students (8-11, 12-15 and 16-18 years old) and three focus groups of teachers according to the level they taught, for each institution. Each group included between 6 and 8 participants. In total, up to 504 participants (half intervention, half control) were projected to participate. Included were teachers over the age of 18 who were employed in 2023-2024 and students between 6 and 18 years of age enrolled in grades 2-11. Non-Spanish speakers or those who did not agree to participate were excluded. Informed consent was obtained from adults and guardians once they were informed of the details, risks and benefits, and consent was obtained from minors.

#### Ethical statements

The study protocol and all procedures were approved by the Institutional Review Boards (IRB) of the XXXXXX on 05/05/2022 (Ref XXXX). All methods were developed in accordance with relevant guidelines and regulations.

## Data Collection

### Variables

Four FGs groups were conducted in each participating educational center: one with teachers and three with students grouped by academic level or age range: one comprising teachers and three consisting of students grouped by academic level or age range. These FGs were conducted at three time points: baseline (pre-intervention), six months after program initiation (post-intervention), and three months later (follow-up). Seventeen trained psychologists and one speech-language pathologist facilitated the sessions using a semi-structured topic guide. This guide was designed to elicit discussion around perceptions, behaviors, and experiences related to resilience, compassion, and the 3Cs strategy (Table 1). In the focus groups with children and adolescents, the question guide was adjusted in Amazonas and Vaupés, since the term “superheroes” was not completely understandable; in Boyacá, the original version was used. All FGs were audio-recorded and transcribed verbatim by a research assistant.

(Table 1. List of guiding questions for focus groups for students and teachers.)

### Intervention

Between June 2023 and August 2024, the “Conmigo, Contigo y Con Todos” (3Cs) strategy was implemented in 28 educational institutions assigned to the intervention group. This strategy, based on third-generation cognitive-behavioral and psychotherapeutic techniques, aims to strengthen resilience, compassion and prosocial behavior, as well as reduce symptoms of depression, anxiety and post-traumatic stress through the development of life skills The research was conducted by implementing the Conmigo, Contigo y Con Todos (3Cs) strategy, which focuses on enhancing resilience, compassion, and prosocial behavior, while also aiming to reduce symptoms of depression, anxiety, and post-traumatic stress using life skills training delivered across 12 program modules (González Ballesteros et al., 2021a).

Teachers from these schools participated in a six-month face-to-face training process, which included learning and appropriation of the 12 modules of the 3Cs program, focused on topics such as self-awareness, empathy, assertive communication, problem solving, emotional regulation, compassion and resilience. Accompanied by local facilitators, the teachers culturally adapted the contents to the context of their school communities with the objective of having a simpler language for the children and adapted to the knowledge that each group had from their region and their daily life. Vaupés and Amazonas were the regions with the greatest need for adaptations, where multiple practical exercises were replaced by their own cultural practices, which were recognized as strategies in which the community collected the topics raised in the model, despite nominating them differently, and also analyzed and incorporated additional topics in the same practices to facilitate their understanding. Even differential adaptations were made for the application, understanding that in the same group of children there were differences in concepts and comprehension based on school years, ethnicity and area of residence between rural and urban regions. Teachers and students, as needed, participated in the process of translation into indigenous dialect when necessary. After that adaptation, teachers replicated the training with one or two groups of students per institution, in six to eight face-to-face sessions held during the regular academic calendar. The children and adolescents in the control group did not receive the intervention during the study and only accessed the 3Cs strategy at the end of the nine-month intervention period.

## Data Analysis

FG transcripts were analyzed using Braun and Clarke’s six-step thematic analysis method (Kiger & Varpio, 2020). This approach facilitated the identification, organization, and interpretation of key patterns and themes related to participants’ perceptions of resilience and compassion, and the changes observed following the intervention (González Gil & Cano Arana, 2010). NVivo14 software was used to support the deductive coding process. A sociologist analyzed the data using a predetermined coding framework encompassing four primary categories: resilience, compassion, inclusion, and the 3Cs Model. However, to examine the perceptions and potential changes in the resilience and compassion capacities of children, adolescents, and teachers, this article focuses exclusively on the categories of resilience and compassion.

An evaluation of thematic saturation in the FGs was conducted at each stage of the study (baseline, post-intervention, and follow-up), disaggregated by department and participant group (teachers and students) to ensure that sufficient and meaningful data were obtained for each topic (Guest et al., 2020).

## Results

In total, 166 FGs were conducted with teachers: 57 at baseline, 55 post-intervention, and 54 at follow-up. Additionally, 667 FGs were conducted with students: 234 at baseline, 219 post-intervention, and 214 at follow-up. All FGs were conducted in person, except for one teacher FG at baseline and three at post-intervention, which were conducted virtually because of access limitations in some regions.

(Graph 1. Flow chart. Qualitative study)

After analyzing 142 teacher FG transcripts (57 baseline, 47 post-intervention, and 38 follow-up) and 429 student FG transcripts (234 baseline, 95 post-intervention, and 100 follow-up), a thematic saturation level of over 91% was reached for both the intervention and control groups, as described in Table 2.

(Table 2. Coded focus groups (transcripts) required to reach data saturation level)

## Resilience Capacity

The data revealed a rich variety of meanings and lived experiences related to resilience among both students and teachers across the three departments. The analysis identified six core themes: concept of resilience, self-awareness, stress tolerance, interpersonal relationships, emotional management, and spirituality.

Among students in the control group across Amazonas, Vaupés, and Boyacá, a progression in their understanding of resilience was observed over the three data collection points. Initially, students associated resilience with mere endurance or dependency on others. Over time, their perspectives evolved to include self-awareness, mutual support, and emotional coping strategies. In Amazonas and Vaupés, spirituality emerged as a key source of resilience, while in Boyacá, students emphasized learning from adversity. Emotional management and interpersonal relationships showed a shift from avoidance to conscious emotional regulation and a recognition of interdependence.

In contrast, teachers in the control group exhibited less change over time. In Vaupés, some teachers remained unfamiliar with the concept of resilience even at the follow-up stage; while in Amazonas and Boyacá, resilience was more commonly linked with adaptation and coping strategies. Teachers consistently emphasized self-reflection, self-compassion, social support, and spirituality across all three regions and time points of the study. Support networks were assessed throughout, although in Vaupés, crises were specifically regarded as opportunities to strengthen social bonds.

In the intervention group, teachers’ perceptions of their own resilience evolved throughout the three measurement stages. Initial definitions focused on overcoming hardship; over time, resilience was regarded as encompassing self-care, empathy, and the capacity to learn from adversity. In Amazonas, the concept of resilience evolved from a family- and spirituality-based understanding to a more structured, individual-centered approach emphasizing patience and fortitude. In Vaupés, teachers described incorporating specific strategies from the 3Cs Model that enhanced their self-regulation and decision-making. In Boyacá, the shift involved a deepened reflection on self-care with the realization that caring for oneself is a prerequisite for supporting others.

Teachers’ reflections on self-awareness advanced from a general identification of strengths and weaknesses to a more nuanced awareness of emotional processes and the value of regulation. In Vaupés and Boyacá, teachers highlighted the 3Cs strategy as instrumental in developing emotional self-regulation skills. Stress tolerance improved across all three departments. Teachers moved from narratives of distress and overload to describing the use of specific strategies such as patience, empathy, and problem-solving to manage difficult situations.

Spirituality remained constant as a key coping mechanism, particularly in Vaupés, where its role as an emotional guide was emphasized more strongly. Emotional management also shifted from an intuitive, unstructured approach to one characterized by a more conscious application of regulation strategies, such as self-care and empathy. Finally, interpersonal relationships were universally recognized as essential for well-being. Teachers across all three departments emphasized the importance of support networks, both professionally and personally.

(Figure 2. Quotes for the concept of resilience and emotional management, intervention teachers)

Among students in Amazonas, Vaupés, and Boyacá, the understanding of resilience evolved significantly over the course of the intervention. Initially, resilience was either unfamiliar to many students or was narrowly associated with enduring adversity. By the third measurement, their conceptualization had broadened to encompass key elements such as empathy, self-reflection, and the ability to derive learning from challenging experiences.

Regarding self-awareness, the early stages of the intervention were marked by frequent expressions of fear and difficulty in identifying and articulating emotions. Over time, students began to demonstrate increased self-motivation and more effective emotional regulation. In Vaupés and Boyacá, several students specifically referenced the 3Cs Model as instrumental in facilitating self-reflection and emotional awareness using collaborative group activities. Similarly, stress tolerance showed noticeable improvement. Initially, students tended to adopt avoidant coping mechanisms such as social withdrawal or distraction. However, by the final measurement, they reported the adoption of emotional regulation techniques and problem-solving strategies. In Vaupés, a specific 3Cs activity was recognized for helping students identify past emotional experiences that continued to influence their present behavior.

Spirituality consistently emerged as an important psychological and cultural resource across the three territories, albeit with regionally distinct expressions. In Amazonas and Vaupés, spirituality was primarily perceived as a source of personal strength and trust, whereas in Boyacá, it played a key role in reinforcing family connections. In the domain of emotional management, students gradually shifted from impulsive or reactive responses to more deliberate emotional regulation, emphasizing the importance of dialogue and self-care. Interpersonal relationships also showed notable development. At baseline, students often described family and peer networks as emotional refuges. By the third measurement, they began to value interpersonal exchanges as opportunities for introspection.

(Figure 3. Quotes for the concept of resilience and interpersonal relationships, intervention students)

A more detailed account of the intervention group’s findings, organized by theme, territory, and measurement point, is presented in Table 3.

(Table 3: Resilient capacity for the intervention group according to participant, measurement, and territory)

## Compassionate Capacity

In relation to compassionate capacity, diverse perceptions and experiences emerged among both students and teachers. These were thematically categorized into three concept areas: compassion, empathy, and self-compassion.

In the control groups from Vaupés, Amazonas, and Boyacá, participants’ understanding of these concepts also evolved though to varying degrees. Initially, students conceptualized compassion as a set of concrete helping behaviors, often devoid of deeper intentionality or reflection. Empathy was largely limited to recognizing others’ discomfort without necessarily engaging with or understanding their emotional experiences. Self-compassion was largely absent from early narratives, often replaced by patterns of self-criticism and reliance on external validation. However, by the third measurement students had developed a more complex view, framing compassion as a shared responsibility and an ethical stance of care. Empathy was redefined as a deeper emotional understanding, while self-compassion emerged as a strategy for personal resilience and collective well-being.

In contrast, teachers demonstrated comparatively less change over time. In Vaupés, some teachers continued to equate compassion with pity, which contributed to emotional exhaustion because of the persistent prioritization of others’ needs. In Amazonas, the concept of compassion expanded to include concern for animals and the natural environment, though the connotation of pity remained. In Boyacá, self-compassion was occasionally acknowledged but lacked the support of practical strategies, leaving teachers dependent on external sources.

Among teachers in the intervention group across Amazonas, Vaupés, and Boyacá, perceptions of compassion, empathy, and self-compassion evolved over the course of the study. Initially, teachers in Amazonas tended to associate compassion with pity or weakness, whereas in Vaupés and Boyacá, it was more often understood as empathy and emotional support. By the post-intervention and follow-up measurements, educators in all three regions demonstrated a significant shift in their understanding of compassion, emphasizing its connection to autonomy, cultural sensitivity, active listening, and accompaniment within their pedagogical practices.

Regarding empathy, teachers in Amazonas and Vaupés underlined the value of dialogue and active listening, although they initially reported challenges in setting emotional boundaries. By the third measurement, they recognized that the 3Cs Model had helped them develop emotional management skills and offer support without compromising their own well-being. In Boyacá, teachers echoed similar themes, acknowledging the importance of boundary setting and referring students to mental health professionals when needed. The spaces created by the 3Cs Model were frequently cited as instrumental in cultivating a more balanced and sustainable approach to empathy and emotional support.

As for self-compassion, teachers across all regions initially found it difficult to prioritize their well-being or effectively manage their emotional responses. However, over the course of the intervention, many described incorporating self-care practices, forgiveness, and seeking support in their personal reflection. In Vaupés and Boyacá, these improvements were directly attributed to the tools and spaces provided by the 3Cs Model.

(Figure 4. Quotes for the concept of compassion and concept of empathy, intervention teachers)

Similarly, students in the intervention group demonstrated significant evolution in their understanding and enactment of compassion-related capacities. In Amazonas, the initial perception of compassion—rooted in pity and sorrow—was gradually redefined to include emotional support, respect, and awareness of others’ suffering. In Vaupés, students shifted from an individualistic approach to a more collective one, emphasizing the value of coexistence, companionship, and forgiveness. In Boyacá, where compassion was initially tied to nostalgia and helping behavior, it transformed into an expression of solidarity and self-love that distinguished itself from pity.

As for empathy, in Amazonas and Vaupés, students moved from associating it with emotional support to recognizing it as nonjudgmental understanding and active accompaniment. In Boyacá, the understanding of empathy shifted from an emotional approach to a more practical one, in which accompaniment, finding solutions, and generating well-being were key elements. Perceptions of self-compassion also changed significantly across the three territories. At the beginning of the intervention, students often described experiences of sadness, emotional hardship, and a dependence on others for support. However, by the third measurement, a stronger vision emerged, highlighting the use of coping strategies, importance of recognizing and learning from mistakes, and central role of resilience in fostering emotional well-being.

(Figure 5. Quotes for the concept of compassion and concept of empathy, intervention students)

A comprehensive analysis of compassionate capacity within the intervention group—disaggregated by theme, territory, and measurement—is presented in Table 4.

(Table 4: Compassionate capacity for the intervention group according to participant, measurement, and territory).

## Discussion

This study aims to examine how perceptions of resilience and compassion evolved among children, adolescents, and their teachers from three departments in Colombia, following the implementation of the 3Cs Model. The findings indicate that resilient and compassionate capacities are not static or uniform traits but rather dynamic constructs shaped by diverse emotional experiences and sociocultural contexts. Significant transformations were observed in the perceptions of both students and teachers within the intervention group. Participants reported that the 3Cs Model facilitated processes of self-reflection, emotional regulation, mutual care, and the re-signification of suffering. In this sense, resilience and compassion were not solely understood as responses to adversity, but as capacities that contribute to the promotion of community processes of emotional regulation and well-being.

These findings align with previous research, such as Amini-Tehrani et al. (2020), which highlights the relational and multifactorial nature of resilience. According to this research, resilience is deeply influenced by the social environment, socioeconomic conditions, and socio-emotional relationships with family and peers. Furthermore, factors such as compassion, self-awareness, and emotional regulation—identified in the present study— (McCrea et al., 2019), underscore the complexity of resilience as a capacity that emerges from the interplay of multiple determinants (Amini-Tehrani et al., 2020; Cicchetti, 2010; González Ballesteros et al., 2021b; Tol et al., 2013b). Consistent with this, our results demonstrate that the development of resilience and compassion varied across territories, educational stakeholders, and the stages of intervention, further affirming their status as context-dependent and evolving constructs.

Teachers’ reflections following the implementation of the 3Cs Model revealed a deepened understanding of both resilience and compassion. They emphasized the value of self-care, empathy, and interpersonal relationships in fostering well-being and strengthening support networks. This is consistent with existing literature on protective factors, which suggests that individual and contextual elements—such as altruistic motivation and positive collegial relationships—serve as buffers against occupational stress. Moreover, awareness and regulation of one’s own emotions, as well as those of others, are widely recognized as key contributors to the development of resilience among educators (Beltman et al., 2011).

Similarly, students across the three departments demonstrated a progressive transformation in their conceptualizations of resilience and compassion. Their evolving understanding encompassed adaptive coping strategies, emotional regulation, and an increased sense of solidarity and mutual support. These changes are consistent with factors highlighted in prior research, such as coping efficacy, self-efficacy, and an internal locus of control (Luthar & Cicchetti, 2000; Tol et al., 2013b). The students’ appropriation of specific strategies for emotional management and relational awareness suggests that the 3Cs Model fosters critical thinking around well-being. This is aligned with principles of health literacy, which emphasize the development of cognitive tools that enhance individual and collective capacities to navigate adversity.

The sustained enrichment of perceptions related to resilience and compassion, observed between the second and third measurement points, indicates a potential consolidation of learning and enduring effects of the 3Cs Model. Previous studies have noted the lack of long-term follow-up as a limitation in evaluating the effectiveness of school-based interventions (Ahmad et al., 2020; Chisholm et al., 2016; Ma, Anderson et al., 2023). Therefore, these findings underscore the importance of incorporating longitudinal and qualitative approaches to fully capture the scope and depth of such initiatives (Navarro de Vasconcellos et al., 2024).

Similar to other interventions implemented in contexts of violence and adversity, school-based MHPSS programs in humanitarian settings have shown potential to strengthen children’s psychosocial well-being and learning. They do so by building coping skills, strengthening interpersonal relationships, increasing social support and feelings of security, and enhancing emotion regulation. These mechanisms, embedded within program theories that also consider the roles of teachers, caregivers, and the broader educational ecosystem, offer a valuable foundation for improving intervention design and implementation in settings affected by adversity (Lasater et al., 2022).

In this regard, the study presents evidence of significant transformations resulting from a school-based community-based psychosocial intervention. As emphasized in contemporary research and endorsed by institutional frameworks, educational settings are key environments for the promotion and prevention of mental health challenges among adolescents and youth (Casañas & Lalucat i Jó, 2018; Comisión de las Comunidades Europeas, 2005; World Health Organization, 2004). Moreover, the dual perspective on resilience and compassion—examined through both teacher and student experiences—reinforces the necessity of intersectoral approaches. Effective mental health promotion requires the engagement not only of health professionals but also of key stakeholders within the socio-educational context such as teachers, who play a vital role in shaping the emotional and social environments of children and adolescents (Casañas & Lalucat i Jó, 2018).

Finally, educational policies should place greater emphasis on empowering students by deliberately cultivating socio-emotional competencies (Mondi et al., 2021). Systematic reviews have consistently demonstrated the effectiveness of frameworks such as the PERMA(H) Model—which encompasses positive emotions, engagement, relationships, and skills such as empathy and gratitude— in enhancing the well-being and positive development of children and adolescents (Benoit & Gabola, 2021). In line with these findings, the results of this study provide a foundation for designing educational programs, modules, or activities that prioritize self-reflection, emotional regulation, mutual care, and the re-signification of suffering as key components of student development. Such programs should move beyond the mere transmission of mental health knowledge to actively foster the emotional and social capacities essential for holistic well-being (Metsäpelto et al., 2010).

Moreover, enabling teachers to personally engage in processes of self-reflection, emotional regulation, and mutual care—as reported through their experience with the 3Cs Model—could significantly enhance the transfer of these skills to students. This approach would enable schools to be addressed as integrated environments, simultaneously strengthening the individual health literacy competencies of both students and teachers, while also fostering the organizational structures and capacity-building processes necessary to sustain and institutionalize such literacy within the educational system (Okan et al., 2020).

A model with strong public policy potential due to its capacity for replication by local community actors.

Since its design and implementation, the 3Cs Model has shown strong potential for community scalability. It’s simple, replicable, and culturally adaptable structure allows it to effectively respond to the specific needs of each territory. The model enables efficient training of local actors—teachers, families, and school networks—who then serve as multipliers in their own communities. This approach has fostered local ownership, even in areas affected by inequality and violence, strengthening community and school networks as protective environments.

In public policy terms, the 3Cs Model represents a viable, effective, and low-cost strategy for promoting socioemotional skills and preventing mental health risks. Its minimal resource requirements make it suitable for low-income settings with limited access to specialized personnel. By encouraging the development of resilient, compassionate, and prosocial behaviors, the model helps build safer, more supportive environments. It also promotes intersectoral collaboration between education, health, social welfare, and community development sectors, enhancing its institutional reach and sustainability. This intersectoral articulation supports its integration into local development plans and public agendas. Through the creation of local capacities and participatory processes, the 3Cs Model not only produces short-term outcomes but also ensures the continuity of actions, positioning itself as a strategic tool for long-term, community-based social transformation.

## Strengths and Limitations of the Study

This study incorporates the perspectives of students and teachers from three departments in Colombia. The findings are enriched by a diversity of experiences and viewpoints on resilience and compassion, shaped by the territorial and cultural context of the participants. These results contribute meaningfully to the broader discussion on the potential of school- and community-based interventions to promote socio-emotional skills, which play a critical role in fostering protective factors associated with well-being and resilience—particularly among students and educators living in conditions of adversity, such as those affected by Colombia’s ongoing armed conflict.

Nonetheless, the findings of this study are inherently tied to the subjective experiences of the participants and specificities of their educational and community contexts. As such, the ability to generalize these results to other vulnerable populations or geographic areas is limited.

## Conclusion

The findings of this study demonstrate that resilience and compassion among teachers and students are multifaceted constructs, influenced by geographic, emotional, and educational variables. In all three departments, participants in the intervention group exhibited notable transformations in their understanding of these concepts, moving from narrow or limited interpretations toward more holistic perspectives that integrate self-care, empathy, emotional regulation, and the reinforcement of support networks.

The *Conmigo, Contigo y Con Todo*s -3Cs- Model was consistently identified by both students and teachers as a meaningful tool for fostering relational processes of shared reflection and collective meaning-making It encouraged the development of coping strategies, emotional awareness, and mutual care. These results underscore the importance of implementing context-sensitive mental health promotion and prevention programs. Such interventions are essential for enhancing individual and community well-being, particularly in environments marked by social vulnerability and adversity. However, questions remain open about the sustainability of these effects in the long term, as well as about the adaptation of the model to other educational and cultural contexts, which raises relevant opportunities for future research.

## Data Availability

All data produced in the present study are available upon reasonable request to the authors

## Study Registration Number

NCT05378412. Application date: 05/12/2022

## Ethical Declarations

The study protocol and all procedures were approved by the Institutional Review Boards (IRB) of Pontificia Universidad Javeriana on 05/05/2022 (Ref. 2022/080). All methods were conducted in accordance with relevant guidelines and regulations.

## Availability of Data and Materials

The data have not been deposited in a public repository. The de-identified dataset will be available upon reasonable request from the corresponding author and subject to a data-sharing agreement.

## Author Contributions

LMGB conceived the study. OEGC and CACR supervised data collection. CACR assessed data saturation. IOJ performed data analysis. IOJ and KAS drafted the manuscript. LMGB and CACR revised the manuscript. LEM and JCM helped with manuscript revisión and edition before submission. All authors read and approved the final version.

## Acknowledgements

The authors would like to express their sincere gratitude to all study participants for generously sharing their data, experiences, and perspectives.

## Non-author Contributions

We would like to thank the professionals and support staff for their contributions to participant recruitment, data collection, and data analysis.

We also thank the teachers, schools, and local Departments of Education for facilitating the implementation of project activities within their educational communities.

Special thanks to Jenny Paola Hoyos Troncoso for her role in coordinating and supporting the project: Effect of the “Conmigo, Contigo y Con todos” Model on Students and Teachers in Vaupés, Amazonas, and Boyacá.

