## Supplementary material for "Building Resilience and Compassion in Conflict-Affected Communities: A School-Based Phenomenological Study in Colombia": Figures and tables

**Figure 1: Flow chart. Qualitative study**.

Focused Schools

(n=56)

Randomized Schools

(n=56)

Recruitment

Baseline

**Operated as intervention**

(n=28 Schools [Amazonas (n=5 schools), Vaupés (n=5 schools, Boyacá (n=18 schools)])

**Focus Groups**

(n=29 FGs Teachers; n=122 FGs NNA)

**Operated as control**

(n=28 schools [Amazonas (n=5 schools), Vaupés (n=5 schools, Boyacá (n=18 schools)])

**Focus Groups**

(n=28 FGs Teachers; n=112 FGs NNA)

Post-intervention

**Intervention**

**Focus groups**

(n= 28 FGs Teachers; n= 110 FGs NNA)

**Control**

**Focus groups**

(n= 27 FGs Teachers; n= 109 FGs NNA)

Follow-up

**Intervention**

**Focus groups**

(n= 28 FGs Teachers; n= 103 FGs NNA)

**Control**

**Focus groups**

(n= 26 FGs Teachers; n= 111 FGs NNA)

**Focus groups not conducted**

(n= 2 FGs Teachers)

**Table 1. List of guiding questions for focus groups for students and teachers.**

| Analysis category | Group | **Questions translated into English** |
| --- | --- | --- |
| Resilient capacity | Students | What does it mean to you to be strong when you face problems or difficult situations?  Imagine you are a superhero or superheroine of resilience:  What powers would you have to face life's challenges?  Have you ever had a bad day or a difficult situation?  How did you feel and what did you do to make yourself feel better? |
|  | Teachers | What do you understand by resilience?  Do you remember a difficult situation you have experienced and how you handled it?  What did you learn from that experience?  What difficulties did you encounter in overcoming this difficult situation? |
| Compassionate capacity | **Students** | When you see someone sad or in trouble, how do you feel and what could you do to help that person?  How do you feel when you manage to help a person who is in trouble?  What things make you feel good about yourself when something doesn't go the way you expected?  What do you think the word compassion means?  How can we show compassion to others at school or at home? |
|  | Teachers | What is the first thing you think of when you hear the word compassion?  What do you do when you see a person in difficulty?  How do you feel when you manage to help a person who is in trouble?  How do you take care of yourselves emotionally? |
| 3Cs Strategy | **Students** | Have you taught them about what emotions are and what to do with them?  Have you participated in any activities or games that have helped them feel calmer when they feel angry or sad? What did they do in that activity that helped you feel better?  Have you heard about a course called Conmigo, Contigo, Con Todo?  What did you understand Conmigo, Contigo, Con Todo to be?  What was the biggest learning experience you had at Conmigo, Contigo, Con Todo? |
|  | Teachers | Have you had activities that train you to manage your emotions and deal with your problems in a better way?  Have you heard about Conmigo, contigo y con todo?  What did you understand about Me, with you and with everything?  What do you think of With Me, with you and with everything?  What did you like about it? |

**Figure 2. Quotes for the concept of resilience and emotional management, intervention teachers**

| Concept of Resilience | Emotional Management |
| --- | --- |
| (…) Is the ability of an educator—or any individual—to effectively resolve difficult situations and transform problems into opportunities for positive outcomes. (Educators, Baseline, Vaupés) | Each situation offers a lesson. In my case, for example (…), I have learned to value small moments more and to be cautious about whom I trust. (Educators, Baseline, Amazonas) |
| This training has enabled us… to better understand ourselves and others, and to reflect on how we can contribute to social transformation, right? I believe resilience should be practiced continuously in the classroom, as educators we must remain adaptive. (Educators, Post-intervention, Vaupés) | So, every time I feel overwhelmed, like out of breath (…) I close my eyes, seek a quiet space, try to remain calm, and take deep breaths (…), before seeking a solution. (Educators, Post-intervention, Amazonas) |
| Is the capacity to face challenges and use those experiences as life lessons that help us deal more effectively with future difficulties (Educators, Post-intervention, Vaupés) | Ultimately, these experiences teach us and help us develop our human side in more fulfilling ways. Even simple gestures—listening to someone or greeting them warmly—can brighten another person's day. (Educators, Follow-up, Amazonas) |

**Figure 3. Quotes for the concept of resilience and interpersonal relationships, intervention students**

| Concept of Resilience | Interpersonal Relationships |
| --- | --- |
| Although I had not previously heard the term, I think it means what my colleague said, kind of resisting adversity. (Student, Baseline, Amazonas) | I often seek out people who can advise, support, and listen to me. (Students, Baseline, Boyacá) |
| Well, I have heard the term resilience before, and I associate it with an innate human capacity—one that involves helping others, showing empathy, demonstrating understanding, and offering support to those in need. (Students, Post-intervention, Amazonas) | Currently, I am going through a difficult time. My mother, friends, cousins, aunts, and psychologist have all provided me with significant emotional support. (Students, Port-intervention, Boyacá) |
| I understand resilience as the human capacity to confront disruptive or challenging situations that affect our lives and to move forward despite them. (Students, Follow-up, Amazonas) | I have learned that surrounding ourselves with positive individuals who give us strength is very beneficial and helps us navigate challenging situations. (Students, Follow-up, Boyacá) |

**Figure 4. Quotes for the concepts of compassion and empathy, intervention teachers.**

| Concept of Compassion | Concept of Empathy |
| --- | --- |
| It is like saying “oh poor thing,” like to pity someone. So, compassion is like saying “have compassion for the child”! (Educators, Baseline, Amazonas) | Well, in the classroom, one sees many children with difficulties and needs, let us put it that way. There is a child, for example, who lives with his grandparents, and often lacks basic supplies. I, at least, always keep pencils, erasers, and sharpeners for everyone. I have them on my seat, ready for them to grab. (Educators, Baseline, Vaupés) |
| I believe compassion involves putting oneself in another’s shoes and feeling what the other person feels, right? Perhaps it is also a bit related to… well to that, to putting oneself in the place of others. (Educators, Post-intervention, Amazonas) | I believe that when people are going through hard times, we must show solidarity, offer guidance, and act with compassion toward everyone. (Educators, Post-intervention, Vaupés) |
| It refers to reaching out to others and helping them—not out of pity, but from a genuine desire to support them. It includes teaching others that they too have a role in overcoming their difficulties. (Educators, Follow-up, Amazonas) | Personally, it was very useful to me, because it helped me see things I previously had not noticed. I did not perceive the way you gave it to us. It made me realize that when I feel good about myself, I am better able to support my students and relate to them with understanding and care. (Educators, Follow-up, Vaupés) |

**Figure 5. Quotes for the concept of compassion and concept of empathy, intervention students.**

| Concept of Compassion | Concept of Empathy |
| --- | --- |
| (…) To feel pity. Let us say, to understand the person who is going through a difficult situation. (Students, Baseline, Vaupés) | I believe that the most important qualities of a person are their values (…), to be empathetic, and to take others into account, just like the 3Cs Model suggests: with me, with you, and with everyone. (Students, Baseline, Boyacá) |
| Compassion is like feeling that affection or feeling. It is like putting yourself in the shoes of the person who is feeling bad, is it not? It is like offering them support, so that they feel company, they feel that they can count on someone. (Students, Post-intervention, Vaupés) | Somehow, the workshops have helped us confront these problems with maturity and with greater empathy. They taught us to recognize that everyone has challenges and that developing empathy helps us face adversity and manage our emotions. (Students, Post-intervention, Boyacá) |
| As it was explained in the training module. I still remember that compassion goes beyond empathy. It involves not only understanding others’ pain but also taking action to alleviate it (Students, Follow-up, Vaupés) | Well, it involves protecting ourselves and others, learning how to support ourselves and others. And also, to develop the ability to work both independently and in collaboration with others. (Students, Follow-up, Boyacá) |

**Table 2: Coded focus groups (transcripts) required to reach data saturation level.**

| **Thematic saturation** | | **Teachers** | | | | | | | **Students** | | | | | | | | |
| --- | --- | --- | --- | --- | --- | --- | --- | --- | --- | --- | --- | --- | --- | --- | --- | --- | --- |
|  |  | **Baseline** | | **Post-intervention** | | **Follow-up** | | | **Baseline** | | | **Post-intervention** | | | **Follow-up** | | |
| **Department** | **Description** | Intervention Group | Control Group | Intervention Group | Control Group | Intervention Group | Control Group | | Intervention Group | | Control Group | Intervention Group | | Control Group | Intervention Group | Control Group | |
| **Vaupés** | **Number of transcriptions** | 4+2 | 3+2 | 3+2 | 3+1 | 3+2 | 3+1 | | 4+2 | | 3+2 | 14+2 | | 11+2 | 7+2 | 5+2 | |
|  | **% saturation** | 100% | 100% | 100% | 100% | 84%* | 100% | | 100% | | 100% | 100% | | 100% | 100% | 100% | |
| **Amazonas** | **Number of transcriptions** | 3+2 | 3+2 | 3+2 | 3+2 | 3+2 | 3+2 | | 3+2 | | 3+2 | 3+2 | | 3+2 | 8+2 | 7+2 | |
|  | **% saturation** | 89%* | 86%* | 100% | 100% | 90%* | 95% | | 89%* | | 86%* | 100% | | 100% | 100% | 100% | |
| **Boyacá** | **Number of transcriptions** | 12+2 | 10+2 | 7+2 | 6+2 | 5+2 | 3+2 | | 10+2 | | 12+2 | 7+2 | | 6+2 | 14+2 | 11+2 | |
|  | **% saturation** | 100% | 94%* | 100% | 100% | 100% | 100% | | 94%* | | 100% | 100% | | 100% | 100% | 100% | |
| **Subtotal group** | | **96%** | **93%** | **100%** | **100%** | **91%** | | **98%** | | **94%** | **95%** | | **100%** | **100%** | **100%** | | **100%** |
| * Saturation of more than 95% is not achieved, as no further information is available for coding. | | | | | | | | | | | | | | | | | |

**Table 3: Resilient capacity for the intervention group according to participant, measurement, and territory**

| **Topic** | **Participant type** | **Measurement** | **Amazonas** | **Vaupés** | **Boyacá** |
| --- | --- | --- | --- | --- | --- |
| **Resilience concept** | **Teachers** | **Baseline** | They articulate a broad, collective understanding of resilience, in which family support, spirituality, and the capacity to cope with change and adversity play a central role. | They understand resilience as the ability to recover from, adapt to, and actively confront life’s difficulties, with particular emphasis on perseverance and the transformation of negative experiences into constructive outcomes. | They associate resilience with problem-solving skills, the capacity to embrace change, and the inclination to support others—whether emotionally or through material means. |
|  |  | **Post-intervention** | Subtle distinctions emerge, linking resilience to attitudes such as courage, inner strength, and patience. | They report conceptualizing resilience, based on their experience with the 3Cs Model, as the capacity to navigate adversity and emerge strengthened. | They emphasize insights gained through the 3Cs Model, particularly regarding self-care, noting that one must attend to personal well-being before being able to empathize with and assist others effectively. |
|  |  | **Follow-up** | They consolidate their conceptualization of resilience as a multidimensional quality that enables individuals to maintain emotional balance, pursue goals, and transform adverse experiences into opportunities for meaningful growth. |  |  |
|  | **Students** | **Baseline** | Some individuals express unfamiliarity with the term “resilience” or associate it primarily with enduring hardship, demonstrating bravery, attending therapy, or struggling to express emotions. | Many students are initially unfamiliar with the term “resilience,” and those who reference it tend to associate it with confronting problems, enduring hardship, reflecting on personal identity, and overcoming obstacles. | Some acknowledge that they had never encountered the term “resilience,” while others associate it with enduring emotional pain, withholding judgment, and understanding the experiences of others. |
|  |  | **Post-intervention** | Their understanding broadens to include the ability to overcome challenges, adapt to new circumstances, and demonstrate empathy—often linking resilience to familial contexts and prosocial behaviors such as helping others. | They link resilience to social coexistence, highlighting the relevance of tolerance, respect, and adaptability in the face of challenges. | They describe resilience as the capacity to navigate difficult moments by seeking solutions, requesting support, remaining calm, and confronting challenges with determination to move forward. |
|  |  | **Follow-up** | While some confusion persists—such as misinterpreting the term as referring to a physical place—the dominant perception frames resilience as an internal strength that enables individuals to face adversity and progress. | They mention associating resilience with empathy or caring for the environment, highlighting the importance of “putting oneself in someone else's shoes.” | They identify new dimensions of resilience, including the ability to adopt another’s perspective, adjust to life changes, extract lessons from adversity, engage in reflective thought, and find positivity in the face of challenges. |
| **Self-awareness** | **Teachers** | **Baseline** | Across all three measurement points, they reported having developed the capacity to analyze their emotional responses in challenging situations—such as grief or separation—while some acknowledged continued difficulty in identifying their emotions and behavioral patterns. Nonetheless, these events served as catalysts for introspection, enabling them to recognize personal strengths, limitations, and their capacity to overcome adversity. | They demonstrated increased awareness of their personal strengths and weaknesses. | They underscore the importance of self-awareness and emotional well-being, particularly in the aftermath of challenging experiences such as the pandemic. |
|  |  | **Post-intervention** |  | Statements more explicitly tied to emotional self-regulation emerged, with particular reference to strategies acquired through the 3Cs Model that supported their capacity to manage emotions, accept change, and make more deliberate, effective decision. | They identify the need to engage in emotional reflection and reassess personal priorities as key strategies for enhancing their overall well-being. |
|  |  | **Follow-up** |  |  | They emphasize that the reflective process facilitated by the 3Cs Model enabled them to achieve a better balance between professional responsibilities and personal self-care. |
|  | **Students** | **Baseline** | They expressed experiencing fear in academic settings and noted the importance of seeking distance or support during emotional distress. Simultaneously, they recognized the value of reflection, learning from lived experiences, and appreciating their personal growth. | Reflections surfaced around unresolved emotional losses, feelings of fear and sadness, and the essential role of family support in navigating these experiences. | Their reflections on adverse life experiences include insights into alternative coping strategies they would now consider—such as allowing themselves to cry, taking space from stressors, seeking support through conversation, attending therapy, writing, or actively reframing challenges to find positive aspects. |
|  |  | **Post-intervention** | Although feelings of mistrust were mentioned, they also reported progress in maintaining calm, exercising self-control, behaving more thoughtfully, and placing greater value on familial relationships. | Reflections on the 3Cs Model emphasized how interactive activities and games enabled them to recognize their own capabilities and skills, particularly through the affirmations and perspectives offered by peers. | They acknowledge the 3Cs Model and emotions workshop as valuable tools that enhanced their understanding of emotional processes and responses. |
|  |  | **Follow-up** | They highlighted key insights from the 3Cs Model sessions, particularly the importance of recognizing the struggles of others and cultivating empathy. | They expressed greater self-motivation, improved emotional regulation (particularly regarding anger), an enhanced ability to set personal boundaries, and a growing sense of responsibility. | They report that the 3Cs Model equipped them with concrete strategies to manage emotions more effectively, and they highlight newly acquired practices such as cultivating tolerance, practicing acceptance, focusing on problem-solving, and drawing upon spirituality. |
| **Stress tolerance** | **Teachers** | **Baseline** | They report experiencing feelings of anguish, helplessness, and emotional overload, indicating ongoing challenges in effectively managing stress. | Across all three assessment points, narratives demonstrate varying levels of stress tolerance, including accounts of personal and professional adversity that have contributed to the strengthening of resilience, emotional self-regulation, and adaptive capacity. Nonetheless, some teachers report ongoing difficulties related to uncertainty and occupational stress. | Their capacity for stress tolerance is closely linked to external and internal support systems, including family and social networks, spirituality, self-care practices, physical exercise, and access to psychological therapy. |
|  |  | **Post-intervention** | Their reflections reveal increased awareness of emotional regulation, emphasizing the use of strategies such as patience, empathy, and a more composed approach to navigating challenges. |  | They highlight the 3Cs Model as a fundamental tool in supporting calmer emotional responses. |
|  |  | **Follow-up** |  |  |  |
|  | **Students** | **Baseline** | In the face of difficult circumstances, they describe coping by seeking solutions with calmness and patience, or engaging in distraction strategies like listening to music or participating in activities. Some individuals continue to struggle with unresolved grief, such as the loss of a loved one. | They reflect on learning to be more resilient, adaptive, and capable of processing difficult experiences, though some admit to being significantly affected and express the need for additional support. | Narratives reveal a range of coping responses to difficult situations, such as crying, temporary withdrawal, persistent effort, reframing mistakes, acceptance, and seeking conversations for support. |
|  |  | **Post-intervention** | They acknowledge the challenges inherent in confronting adversity, while also recognizing the importance of acceptance, adaptability, and gratitude as essential components for personal growth and forward movement. | Their reflections on loss and academic challenges are translated into practical strategies, including reframing situations positively, practicing acceptance, and cultivating patience. | The 3Cs Model is recognized for contributing to the development of empathy, maturity in problem-solving, and more conscious emotional awareness. |
|  |  | **Follow-up** | They highlight the value of open dialogue, the ability to learn from adverse experiences, and the development of problem-solving skills. | Stress management strategies emphasize both proactive engagement and emotional self-regulation, with examples such as using physical activity (e.g., boxing) to release anger, avoiding rumination, and expressing emotions constructively. | Their responses emphasize the importance of acceptance, intentional self-care, and professional mental health support (e.g., psychological therapy). |
| **Spirituality** | **Teachers** | **Baseline** | From the initial to the third assessment, spirituality consistently emerged as a core coping resource. Although no substantial shifts were observed in how teachers understood or utilized their spiritual or religious beliefs, many emphasized specific practices—such as prayer, meditation, Bible reading, and participation in the Eucharist. | Across all three data collection points, expressions of faith and spirituality served as both emotional refuge and motivational force. Participants noted that their beliefs offered clarity and purpose in the face of fatigue or stress—anchored in the conviction that challenges occur for a reason and can be endured through trust in a divine plan.  . | Through spiritual practices such as prayer, attending Mass, and engaging with religious texts, they reported accessing feelings of peace, comfort, and mental fortitude. Nevertheless, despite the depth of spiritual engagement, there was limited evidence across the assessments of participants integrating mental health practices as complementary to their spiritual frameworks. |
|  |  | **Post-intervention** |  |  |  |
|  |  | **Follow-up** |  |  |  |
|  | **Students** | **Baseline** | They consistently identified spirituality as a critical support mechanism in the face of hardship, frequently referencing faith in God as particularly meaningful within the context of their Indigenous communities. | Throughout the study, they consistently framed spirituality as a resource for coping, personal transformation, and a foundation for hope and confidence. Faith-based practices—including prayer, religious rituals, and cultural expressions—were reported as vital for fostering emotional and social well-being, offering resilience and a sense of stability amid adversity. Trust in God was repeatedly cited as a fundamental pillar of emotional strength. | In all three assessments, spirituality—particularly expressions of faith in God and reliance on prayer—was viewed as an essential coping strategy, providing strength, emotional balance, and hope in times of hardship. Participants also noted that spirituality extended beyond the individual, strengthening family relationships. However, some mentioned supplementing their coping strategies with non-spiritual methods such as leisure activities, expressive writing, or emotional release through crying. |
|  |  | **Post-intervention** | Their descriptions of spirituality often reflect a moral and behavioral framework rooted in faith, wherein actions are interpreted through religious values and understood as guided or protected by a higher power. |  |  |
|  |  | **Follow-up** |  |  |  |
| **Self-regulation** | **Teachers** | **Baseline** | They emphasize their ability to engage in self-reflection as a mechanism for identifying and implementing solutions to personal and contextual challenges. | They expressed a variety of strategies, such as self-reflection, self-control, and social support to manage emotions, though some acknowledged ongoing challenges in consistently regulating emotional responses. | They identify self-reflection, social support, and physical activity as key methods for managing adverse emotional states. |
|  |  | **Post-intervention** | They report a strengthening of emotional self-regulation capacities through practices such as empathy and self-care, underscoring the role of positive introspection in managing emotions. | Their narratives suggest improvements in emotional self-regulation, albeit with varied strategies adopted by individuals depending on their circumstances and personal development. | They indicate improvement in emotional regulation through the application of self-regulatory strategies, such as boundary setting and tools introduced by the 3Cs Model. |
|  |  | **Follow-up** | They demonstrate a sustained ability to reflect on their experiences and manage emotional responses effectively. |  |  |
|  | **Students** | **Baseline** | They mention strategies such as distancing themselves from emotionally triggering situations or seeking support from trusted individuals as a way to achieve emotional relief. Additionally, coping techniques like using mobile devices or engaging in sports were referenced. | They describe using techniques such as establishing boundaries, emotional distancing, crying, seeking companionship, and self-motivation. | They mention various strategies for managing their emotions, such as crying, venting, distracting themselves, reading, walking, solitude, and seeking psychological therapy. |
|  |  | **Post-intervention** | They describe various strategies for managing emotionally challenging situations, including staying calm during decision-making, requesting support, meditating, talking with others, and spending time in nature—although some continue to experience difficulties with emotional regulation in specific contexts. | They refer to a specific activity within the 3Cs Model that facilitated reflection on previously overlooked experiences that shaped their emotional responses and thought patterns. | While many strategies remain consistent, there is increased reflection on the significance of confiding in trusted individuals, using music as an emotional outlet, and allowing themselves space to process difficult emotions. |
|  |  | **Follow-up** | They refer to therapeutic support, emotional expression, interpersonal assistance, and calm, reflective problem-solving. These practices are presented as outcomes of insights gained through lived experience and intervention activities. | They report employing strategies such as releasing anger physically, managing frustration, avoiding overthinking, and seeking psychological support or dialogue—while also recognizing the emotional benefits of engaging in sports. | New coping mechanisms have emerged, such as turning to spirituality, engaging in video games as a distraction, and intentionally distancing oneself from emotionally taxing situations. |
| **Interpersonal Relationships** | **Teachers** | **Baseline** | They emphasize the vital role of close support networks—including family, friends, and colleagues—in managing emotional challenges and navigating adversity. | Across all three assessments, participants referenced the significance of emotional support from family, friends, neighbors, and coworkers. They noted that such networks provide not only comfort and encouragement but also new perspectives, nonjudgmental listening, and, at times, tangible solutions to problems. | They point to recreational activities and social gatherings as valuable resources for strengthening interpersonal relationships and fostering emotionally safe environments grounded in resilience, trust, and mutual support. |
|  |  | **Post-intervention** | They highlight how relationships with loved ones serve as a central source of motivation and emotional strength, enabling them to overcome difficulties and pursue personal and professional goals. This emotional support is viewed as a key factor in fostering resilience and growth. |  | They affirm that support networks function not only as emotional anchors but also as catalysts for reflection and learning. |
|  |  | **Follow-up** |  |  |  |
|  | **Students** | **Baseline** | They consistently emphasize the importance of support networks—comprising family members, peers, and classmates—in safeguarding emotional well-being and assisting in decision-making during challenging circumstances. | They associate effective coping with interpersonal behaviors such as seeking support from friends and family or consulting mental health professionals, emphasizing relational connections. | The importance of family and social support is consistently reinforced, with particular reference to the emotional and relational benefits of recreational activities and the presence of friends and loved ones. |
|  |  | **Post-intervention** | They acknowledge the positive influence of various coping strategies, including leisure activities, professional mental health support, and reflective practices, which together enhance their capacity to face adversity. | They highlight the interrelationship between resilience, family support, and introspection. While some individuals prefer solitude as a space for emotional processing, although they also engage in music or sports. | There are reflections that emphasize a balance between personal introspection and the active pursuit of external support when navigating complex emotional experiences. |
|  |  | **Follow-up** |  |  |  |

**Table 4: Compassionate capacity for the intervention group according to participant, measurement and territory.**

| **Topic** | **Participant Type** | **Measurement** | **Amazonas** | **Vaupés** | **Boyacá** |
| --- | --- | --- | --- | --- | --- |
| **Concept of Compassion** | **Teachers** | **Baseline** | An ambivalent understanding of compassion is evident: while some participants equate it with pity or emotional weakness, others perceive it as a moral responsibility to alleviate the suffering of others. | They express recognizing compassion as closely aligned with empathy—particularly active listening and the provision of emotional or material support during times of hardship. | They relate compassion to empathy, emphasizing the ability to put oneself in another's shoes to understand their difficulties. Some claim to view it not only as an attitude, but also as a moral or spiritual mandate. |
|  |  | **Post-intervention** | This perspective is gradually evolving, with educators increasingly distinguishing compassion from pity, emphasizing that genuine compassion should empower others rather than foster dependency. | A notable shift emerged in participants’ discourse, moving from basic definitions to deeper reflections that incorporate cultural and ethnic sensitivity, and a view of compassion as an evolving quality shaped by continued learning and self-awareness. | Several participants noted that self-care is an essential component of compassion. |
|  |  | **Follow-up** |  |  | Deeper insights emerged in connection to both their social support networks and educational practices, highlighting the relevance of active listening and emotional accompaniment. |
|  | **Students** | **Baseline** | They associate compassion with feeling sympathy, providing help, and offering forgiveness, although some acknowledged unfamiliarity with the term or uncertainty about its meaning. | Compassion is described through relational and affective behaviors such as caring, accompanying others, understanding, and empathetically engaging—although for some, it still evokes notions of guilt, sadness, or pity. | They associate compassion with actions such as empathizing, offering comfort, providing assistance, forgiving, and demonstrating kindness. |
|  |  | **Post-intervention** | Perceptions of tolerance, generosity, and emotional support expanded, while associations with pity and shame persisted. | Their conceptualization is moving toward a more collective understanding, emphasizing values like empathy, solidarity, respect, kindness, and the importance of shared experiences and coexistence. | They add that compassion includes both grand and modest gestures, with examples ranging from offering advice to providing material support. Participants viewed compassion as a means to cultivate their better selves. |
|  |  | **Follow-up** | They referenced the 3Cs Model as a catalyst for conceptual change, noting that the intervention fostered greater understanding of others’ suffering, reduced resentment, and emphasized respect and supportive behaviors. | Compassion is expressed as an integration of care, forgiveness, collaboration, and empathy, with frequent emphasis on the metaphor of “putting oneself in the other’s shoes.” | They reinforce the idea of compassion as encompassing supportive and altruistic behaviors, while integrating self-love, care for others, and an intentional differentiation between compassion and pity. |
| **Concept of Empathy** | **Teachers** | **Baseline** | They reflect empathic behaviors in their experiences, particularly through emotional support provided via dialogue and active listening. However, some noted initial challenges, such as difficulty in setting personal boundaries and a lack of knowledge on how best to offer support. | They emphasize the significance of active listening and open dialogue in recognizing the emotional needs and suffering of others, often accompanied by gestures of motivation and encouragement. | They recognize empathy as the ability to understand and share in others’ suffering, facilitated by active listening, emotional companionship, and thoughtful advice. |
|  |  | **Post-intervention** | Across both the baseline and follow-up, educators reported that the 3Cs Model significantly strengthened their emotional intervention skills. This development not only enhanced their capacity to support others but also fostered greater self-awareness and emotional reflection. | In both the baseline and follow-up, there was greater evidence of introspection and a strong intent to apply the principles learned from the 3Cs Model in their interpersonal relationships and professional contexts, particularly in the classroom. | Across assessments, educators increasingly acknowledged the importance of setting emotional boundaries in their roles. They credited the 3Cs Model’s dedicated spaces for promoting self-compassion, empathy, and emotional resilience. |
|  |  | **Follow-up** |  |  |  |
|  | **Students** | **Baseline** | They recognize empathy as commonly associated with offering support, initiating conversations, and comforting others through actions such as hugging, motivating, and expressing affection. | While empathy was primarily defined in terms of helping, motivating, and advising, some still equated it with feeling sorry for others. | They further associated empathy with regulating emotions, offering advice, and fostering meaningful emotional connections. Participants referenced the framework of the 3Cs—“with me, with you, and with everyone”—to emphasize relational interconnectedness." |
|  |  | **Post-intervention** | The responses focused on providing help, practicing active listening, and offering material support, with specific reference to the 3Cs Model as instrumental in fostering a sense of equality and mutual care. | They emphasize the notion of empathy expanded to include sensing and understanding another’s emotional state, offering comfort and companionship, and striving for interpersonal harmony. | They add that empathy also involves listening, understanding, conveying love and security, and being nonjudgmental—attributes participants linked directly to the insights gained through the 3Cs Model. |
|  |  | **Follow-up** | A deeper understanding of empathy emerged through expressions such as “putting oneself in another’s shoes,” demonstrating nonjudgmental understanding, and showing solidarity. The intervention was seen as equipping them with practical tools to better support others. | They mention specific empathic actions such as listening, hugging, accompanying someone, and collaboratively seeking solutions—reflecting a shift toward more engaged and proactive forms of empathy. | They emphasize empathy as a form of active accompaniment: being present for others, helping them find solutions, offering comfort and counsel, and ensuring no one feels alone. |
| **Concept of Self-Compassion** | **Teachers** | **Baseline** | They reported initial difficulties in managing their emotions, including expressions of fear, self-judgment, and in some cases, self-harm. These early reflections highlighted a lack of emotional self-understanding and a tendency to be overly critical toward themselves and others. | Some relate self-compassion with calmly facing complex or painful personal experiences, indicating that past hardships continue to shape their emotional lives and coping strategies. | Some express struggle with self-pity. |
|  |  | **Post-intervention** | Across both the baseline and follow-up, they expressed having learned to turn challenges into learning opportunities, strengthening their self-esteem and the intentional prioritization of their own well-being. | The starting line and follow-up focus on prioritizing well-being through activities such as meditation and contact with nature. | They credited the 3Cs Model intervention for helping them incorporate self-reflection, forgiveness, and help-seeking into their understanding of self-compassion. |
|  |  | **Follow-up** |  |  |  |
|  | **Students** | **Baseline** | Unresolved emotional experiences—such as loss, fear, sadness, and anxiety—were common themes, with participants often emphasizing the importance of relying on family and trusted support systems to navigate these difficulties. | Self-compassion is felt through difficult times, with expressions like “I wanted to be alone" or "it was difficult,” as well as the recognition that sometimes “we make mistakes” or “we endure painful situations.” | They mention emotional discomfort included venting to others, intentionally redirecting thoughts toward positivity, walking, solitude, and seeking support. |
|  |  | **Post-intervention** | A more wellness-oriented perspective emerges, with the idea of navigating forward through challenges by acknowledging the need for solitude, crying, and cultivating positive thoughts as essential tools for overcoming difficult times. | The expressions are explicitly linking self-compassion to problem-solving and greater empathy toward others. | They further investigate identifying their emotions, recognizing anger and the need to be heard. They reported new practices like setting boundaries, self-forgiveness, and patience. |
|  |  | **Follow-up** | Common phrases such as “even if you fall, you have to get up,” “vent with people you trust,” and “navigate adversity with someone by your side.” |  | The tone of reflection evolved to be more compassionate and learning-oriented, emphasizing growth through difficulty, emotional distancing from conflict, and owning one’s mistakes. |
